# Comparing randomized trial designs to estimate treatment effect in rare diseases with longitudinal models: a simulation study showcased by Autosomal Recessive Cerebellar Ataxias using the SARA score

**DOI:** 10.1101/2025.01.30.25321311

**Authors:** Niels Hendrickx, France Mentré, Alzahra Hamdan, Mats O. Karlsson, Andrew C. Hooker, Andreas Traschütz, Cynthia Gagnon, Rebecca Schüle, ARCA Study Group, EVIDENCE-RND consortium, Matthis Synofzik, Emmanuelle Comets

## Abstract

Parallel designs with an end-of-treatment analysis are commonly used for randomised trials, but they remain challenging to conduct in rare diseases due to small sample size and heterogeneity. Model-based approaches can be more powerful to detect treatment effects. We investigated the performance of longitudinal modelling to evaluate disease-modifying treatments in rare diseases a simulation study, leveraging as showcase a model of the SARA score progression in Autosomal Recessive Cerebellar Ataxia. We compared the power of parallel, crossover and delayed start designs, investigating several trial settings: trial duration (2 or 5 years); disease progression rate (slower or faster); magnitude of residual error (σ =2/σ =0.5); number of patients (100 or 40); method of statistical analysis (longitudinal analysis with non-linear or linear models; standard statistical analysis). In our simulations, using non-linear mixed effect models resulted in higher power than a rich, sparse linear mixed effect model or standard statistical analysis. Parallel and delayed start designs performed better than crossover designs. Our analysis showed that since disease progression is slow and residual variability is high (for the standard clinician-reported outcome), longer durations are needed for power to be greater than 80%, up to 5 years for slower progression and 2 years for faster progression ataxias.

## 1. Introduction

Randomized parallel designs are the gold standard used for clinical trials, and the statistical analysis is then usually a comparison of change from baseline or end of treatment measurements ^1^. Rare diseases however involve small target populations with heterogeneous presentation of the condition, progression and severity of symptoms, or response to treatment. Other designs may then be considered such as single arm trials, or designs using natural history data, such as patient registries, to inform the design or to use as an external control ^2^. Furthermore, analysing longitudinal data using linear or non-linear mixed effect models has been shown to improve the power of those trials compared to standard approaches ^3,4^.

Autosomal recessive Cerebellar Ataxias (ARCA) are a group of ultra-rare, progressive neurodegenerative disorders. They mainly affect the cerebellum but also induce other multi-systemic damage to other neurological systems, causing impairment to gait, balance, speech and fine motor movements ^5^. More than 100 genotypes have been identified ^5^ and symptoms usually emerge during childhood or early adulthood. ARCAs are prime candidates for targeted molecular therapies or gene therapies due to their genetic etiology. While there is currently no approved disease-modifying treatment for most ARCAs, multiple clinical trials are ongoing, which aim to quantify the treatment effect of such therapies. However, the settings of those clinical trials can be very different depending on the genotype or kind of drugs, and there are still statistical challenges to design randomised clinical trials for these diseases ^6^. Among ARCA genotypes, disease severity at onset and disease progression rate can vary ^7–9^, which have an influence on the designs. Other randomized trial designs have been explored for neurodegenerative diseases. For example the FDA just approved a drug for Niemann-Pick disease type C ^10^, where a crossover design was used to demonstrate the effect of the compound. Delayed start designs are also being investigated to show the effect of disease-modifying drugs for instance in Parkinson’s and Alzheimer’s disease ^11,12^.

Crossover and delayed start designs could be used to account for individual progression rates, but modelling approaches are needed to account for temporal change in the outcome or carryover effect. Due to the genetic diversity of this disease, there are currently no universally applicable biomarkers, and clinician-reported composite scores such as the Scale for Assessment for the Rating of Ataxia (SARA) ^13^ are commonly used as endpoints.

In this work, we propose to explore the advantage of model-based analyses of trials compared to more standard statistical analyses through a simulation framework. We evaluate several clinical trial designs that could be used to quantify a disease-modifying treatment effect according to their power and type 1 error. We simulated data using a longitudinal non-linear mixed effect model (NLMEM) ^14^ that we built to describe – as a showcase example for our methodological approach - the SARA score over the time since onset of symptoms in patients included in the ARCA registry ^15^ and diagnosed with Autosomal Recessive Spastic Ataxia of Charlevoix Saguenay (ARSACS), introducing a hypothetical treatment effect slowing down the disease progression rate. We explored two disease progression rates, a “slow progression rate” and a “fast progression rate” to reflect the diversity of progression rates more generally in ARCAs and to assess its impact on clinical trial design. We also explored other clinical settings of interest, such as the duration of the trial, the number of patients and the magnitude of residual error.

## 2. Methods

We performed a simulation study to evaluate and compare the performance of several clinical trial designs for ARCAs in order to quantify a disease-modifying treatment effect. We investigated several settings, such as the design, the estimation model, the number of patients, the magnitude of residual error and the duration of the trial.

### 2.1 Simulation model

Data was simulated using a previously published NLMEM ^14^ describing the evolution of the total SARA score for an exemplary specific subgroup of patients in the ARCA registry ^15^, namely patients with a genetically confirmed diagnosis of Autosomal Recessive Spastic Ataxia of Charlevoix Saguenay (ARSACS) ^16^. The *j^th^* total SARA score *y_ij_* observed at time *t_ij_* since symptoms onset in individual *i* was described using the following equations:

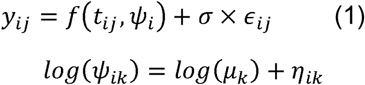

where *f(t_ij_, ψ_i_)* represents the structural model, a function of *t_ij_, ψ_i_*, the vector of individual parameters for individual *i* (of dimension K), *µ* the vector of fixed effects. The random effects η_i_ were assumed to follow a joint multinormal distribution with variance-covariance matrix Ω. For the residual error model, σ was the standard deviation of the residual error, and *∈_ij_ ∼ N(0,1)*. The population parameters for this model were θ = (µ, Ω, σ).

The structural model for the evolution of the SARA score, f was a four-parameter logistic model ^14^:

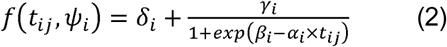

where *ψ_i_ = (δ_i_, γ_i_, β_i_, α_i_)*,

A Drug Effect (DE) parameter was introduced on α, reducing the disease progression rate after the inclusion in the clinical trial in the treatment group, in order to mimic a disease-modifying treatment effect. Therefore, under treatment, we assumed *α_trt_ = α(1 - DE × TRT)*, with *TRT* (treatment) an indicator function (=1 if the patient is treated, 0 if the patient is in the control group). Hence, with a drug effect of 0, the equation is equal to (2), which is equivalent to natural progression with no drug, with a DE of 1, there is no progression, and with DE>1, the SARA score decreases. In this study we assumed that the treatment effect is DE=0.5 hence dividing by 2 the disease progression rate. The parameters used for the simulation are given in Table 1.

**Table 1.** Parameters of the logistic model on the ARSACS population on the ARCA registry used in the simulation study for the slow progression scenario in the control (untreated) group. For the fast progression scenario, α was set to 0.22 and for the low standard error σ was set to 0.5.

| Parameter | Value |
| --- | --- |
| $\delta$ | 6.16 |
| $\gamma$ | 28.75 |
| $\beta$ | 3.94 |
| $\alpha$ (yr<sup>-1</sup>) | 0.11 |
| $\omega_{\delta}$ | 0.31 |
| $\omega_{\gamma}$ | 0 |
| $\omega_{\beta}$ | 0.20 |
| $\omega_{\alpha}$ | 0.09 |
| $\sigma$ | 2 |

### 2.2 Trial designs

Three clinical trial designs were investigated: 1, parallel design where patients are randomized to the control (untreated) group or treatment (treated) group; 2, cross-over, where patients switch group mid-trial; 3, delayed start, where patients are randomized in control or treatment group, and the patients in the control group receive the treatment at mid-trial, as shown in Figure 1. For each design, N patients were included with an allocation ratio of 1:1, and we generated one observation at inclusion in the trial and then every 6 months for the duration of the trial.

**Figure 1.**
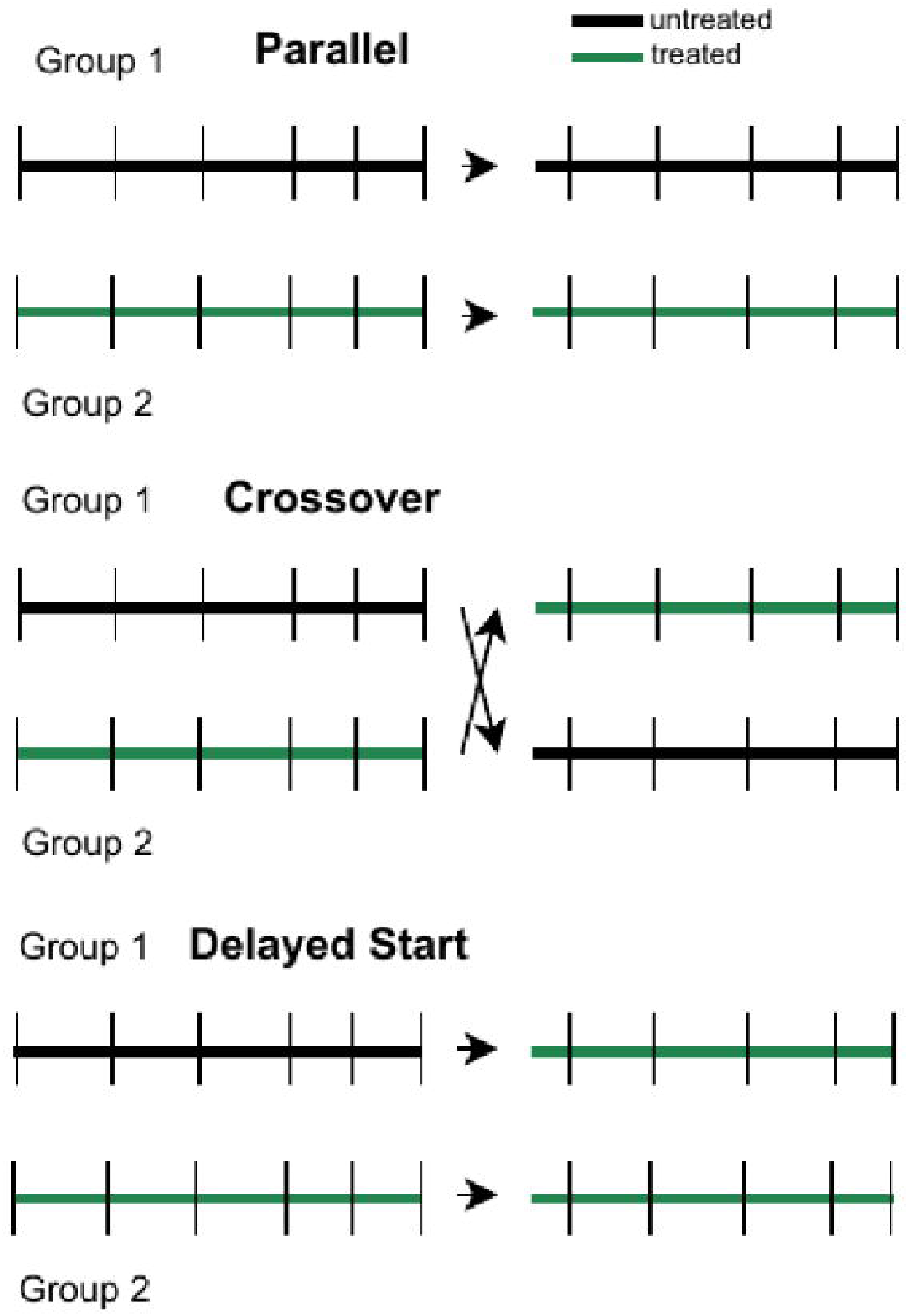
Schematic representation of the parallel, crossover and delayed start designs for a trial, with a change mid-trial in the cross-over and delayed start designs.

### 2.3 Simulation scenarios

We define a simulation scenario as a combination between the parameters of the model used for the simulation, the duration of the trial and the number of patients in the trial. For each scenario, we generated 500 datasets under each of the three trial designs investigated.

In this work, we considered two reference scenarios. The slow progression reference scenario (A) used the parameters previously estimated on the ARSACS population (table 1). In a previous study ^14^, a trial duration of 5 years for the ARSACS population yielded adequate power, so we assumed the same duration.) The fast progression reference scenario (B) assumed a faster disease progression rate than the first, by multiplying the α parameter by 2 (the other parameters remaining unchanged), and we assumed a shorter duration (2 years). The number of observations in each patient is hence 11 for the 5-year trial and 5 for the 2-year trial.

DE was simulated at 0 and 0.5 (for the type 1 error and power assessment, respectively). Figure 2 shows the graph of the logistic model using the population parameters for the slow and fast progression, for treated and untreated patients at three different times since disease onset at inclusion (5, 15 and 25 years).

**Figure 2.**
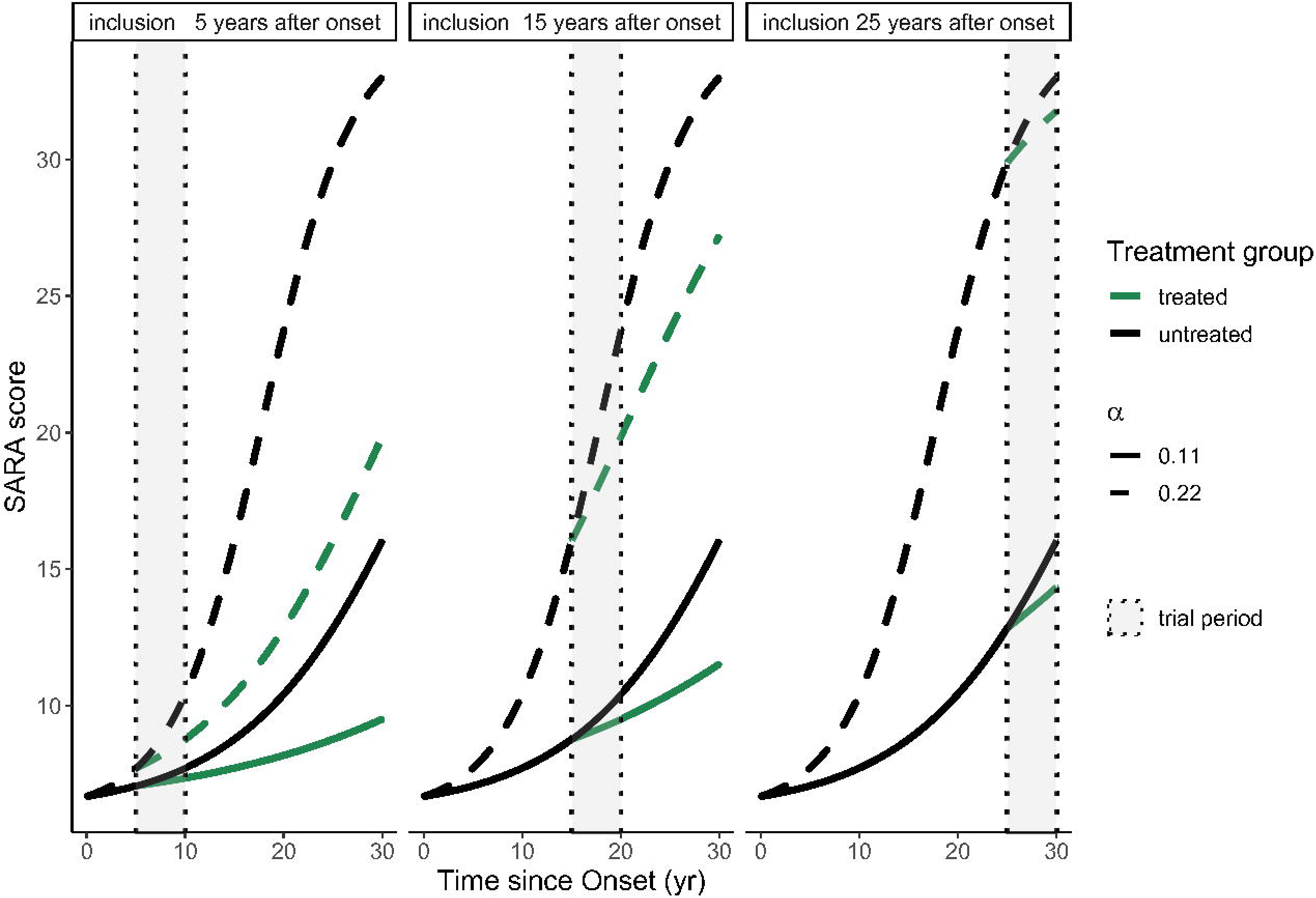
Evolution of Sara score versus time since onset with the logistic model using the population parameters. Each panel represents a time since onset at inclusion (5, 15 and 25 years), full or dashed lines represent two different progression rates (a = 0.11 or 0.22 respectively), the black and green colours represent if the patient is untreated or treated (assuming a drug effect of 0.5), and the grey box represents the trial period.

In each of the reference scenarios, we varied other settings to study the impact on the type 1 error and power of such trials. First, we investigated the influence of the duration of the trial, by using a shorter trial for the slow progression reference (2 years) and a longer trial for the fast progression reference (5 years). We also considered a lower magnitude of the residual error (σ=0.5 with a 2 year and 5-year trial for the slow progression reference, and σ =0.5 with a two-year trial for the fast progression reference).

Time Since Onset (TSO) at inclusion was assumed to follow a uniform distribution between 0 and 30 years ^14^. All scenarios were evaluated with N=100 patients (50 per arm). If, in a scenario, there is at least one design where one model has more than 99% power, all designs and models within that scenario were simulated again with 40 patients. In this additional simulation, the simulated patients were stratified on their time since onset at inclusion to ensure a balanced design (30%: 0-10 years, 30%: 10-20 years, 40%: 20-30 years). The data of the first simulated data set is shown for all designs (slow progression, 5-year duration, 100 patients, with σ=2 and σ=0.5) in Figure 3.

**Figure 3.**
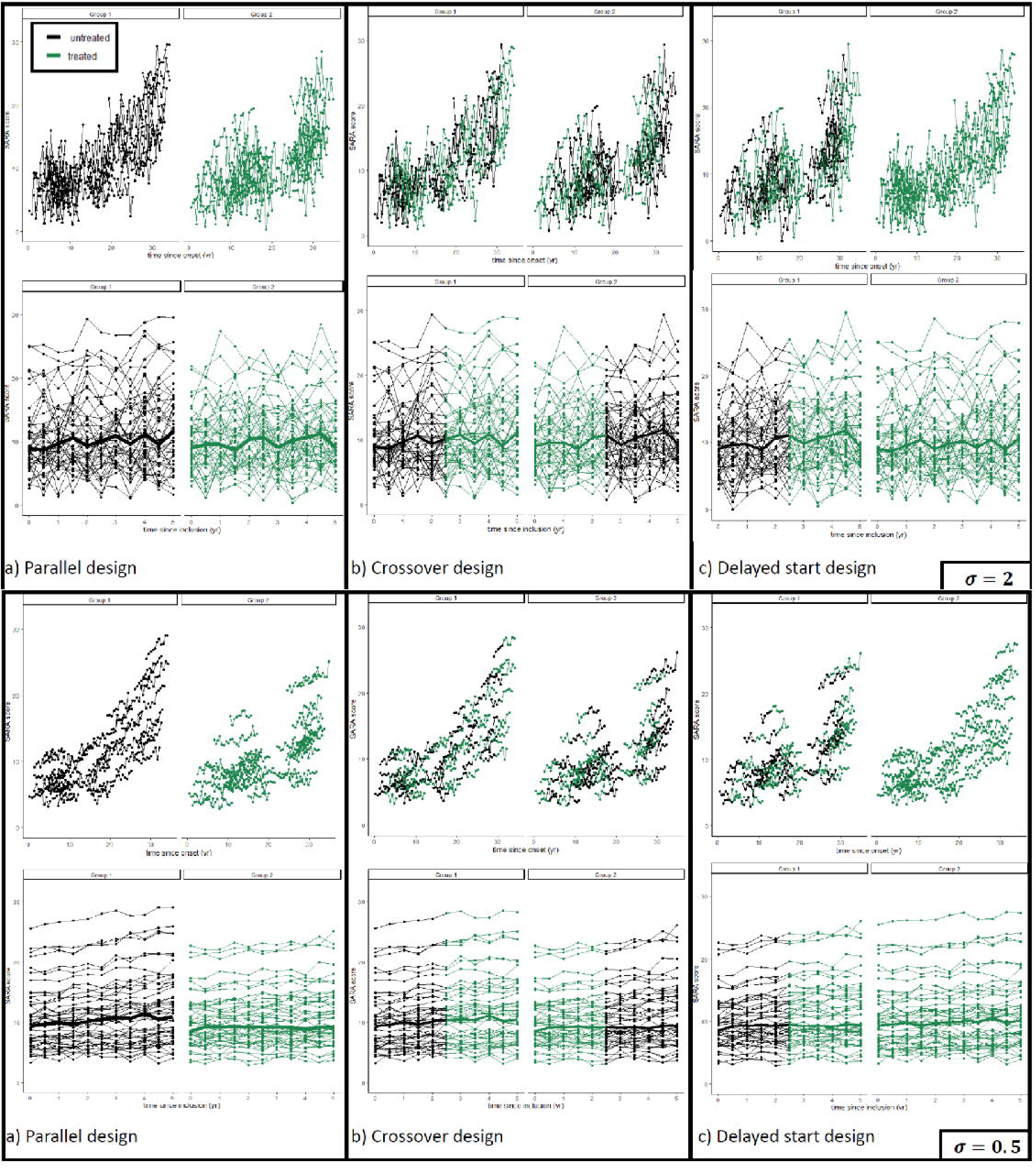
Simulated data set with a parallel (left), crossover (middle) or delayed start (right) design, with a duration of 5 years, N=100 patients. The top two rows represent σ=2, and the bottom two rows σ=0.5, as a function of time since onset of symptoms (top row) or time since inclusion (bottom row). Each panel represents a randomization group, lines join the observations of a patient. Treatment status is differentiated by colour (black: patients that are not under treatment; green: patients under treatment). In the graph versus time since inclusion, the bold line represents the median observation at each time point in each panel.

### 2.4 Estimation models

Each simulated trial was analysed using all observed data with 2 models, the logistic model used for the simulation and a simpler model assuming a linear evolution of the SARA score. For the logistic model, DE was estimated with no random effect and it was fixed to 0 for the crossover and delayed start designs when patients were untreated. Also, to ensure the identifiability of all parameters, γ was fixed to its estimated value in the ARSACS population and its random effect was fixed to 0. For the linear model (with slope and intercept, both with random effects and a covariance), a fixed effect DE was introduced on the slope parameter. Time since inclusion was used as the independent variable, with the time since onset at inclusion as a covariate on the base and slope.

In the slow progression reference scenario, we also assessed the performance of a sparser linear model, which we call sparse linear model. In this approach, we used the same linear model, but only the first, mid-trial and last observations were kept for each patient. It was made to mimic an end-of-study approach, while keeping the mid-trial point, which is needed for the crossover and delayed start designs.

### 2.5 Evaluation

For each scenario, we evaluated the empirical type 1 error of the Likelihood Ratio Test (LRT) comparing a model with and without treatment effect (type I error of 0.05), in a simulation where DE = 0. We also evaluated the power based on the LRT, corrected for the empirical type 1 error (i.e., with a statistical significance threshold chosen such that the type 1 error is 5% at that threshold), by simulating a drug effect of 50% (DE=0.5). For each estimated proportion of significant LRT over the 500 replicates, its 95% confidence interval was constructed using an exact binomial test (using the Pearson-Clopper method).

### 2.6 Implementation

Simulations were performed using NONMEM 7.5.0 ^17^, via the sse command in Pearl Speaks NONMEM (PsN) 5.3.1 ^18^. Parameter estimation was performed using the SAEM ^19^ algorithm for the logistic and linear model, and using the FOCE ^20^ algorithm for the sparse linear model. Data analysis and visualisation were performed using R 4.1.2 ^21^. NONMEM and PsN code for a scenario of the simulation study are available in a Zenodo repository (doi: https://doi.org/10.5281/zenodo.14755801).

## 3. Results

### 3.1 Simulation study: Slow-progression case

Figure 4 shows the type 1 error for the three designs (parallel, crossover and delayed start) and three models (logistic, linear and sparse linear) in the slow progression reference scenario (N= 100, duration 5 years and σ =2). The type 1 error of the logistic model was controlled for the parallel and delayed start design, and slightly inflated for the crossover design. Type 1 error was controlled in every design for the linear model. For the sparse linear model, the type 1 error was controlled for the crossover and parallel designs but inflated for the delayed start design. Figure 5 shows the corrected power for the three designs. For the parallel design, the linear model had lower power than the logistic model (75.4%) and the linear end-of-treatment much lower power (49.2%), whereas the logistic model had the highest power (88%). If we were to apply a two-sample t-test with change from baseline, the type 1 error would be controlled and the power would be even lower than the sparse linear model, at 35.8%. The difference in power for the three designs was even more marked for the delayed start design. For the logistic model, the corrected power was highest for the parallel design (88.0%), closely followed by the delayed start design (83.4%), while the crossover design (44.4%) performed much worse. The two other models followed a similar trend across all three designs. In the supplementary material, Figures S1-1 and S1-2 show the violin plots of the error in the estimation of the treatment effect in the slow and fast progression case, with the nonlinear mixed effect model which were adequate both for DE= 0 and DE = 0.5.

**Figure 4.**
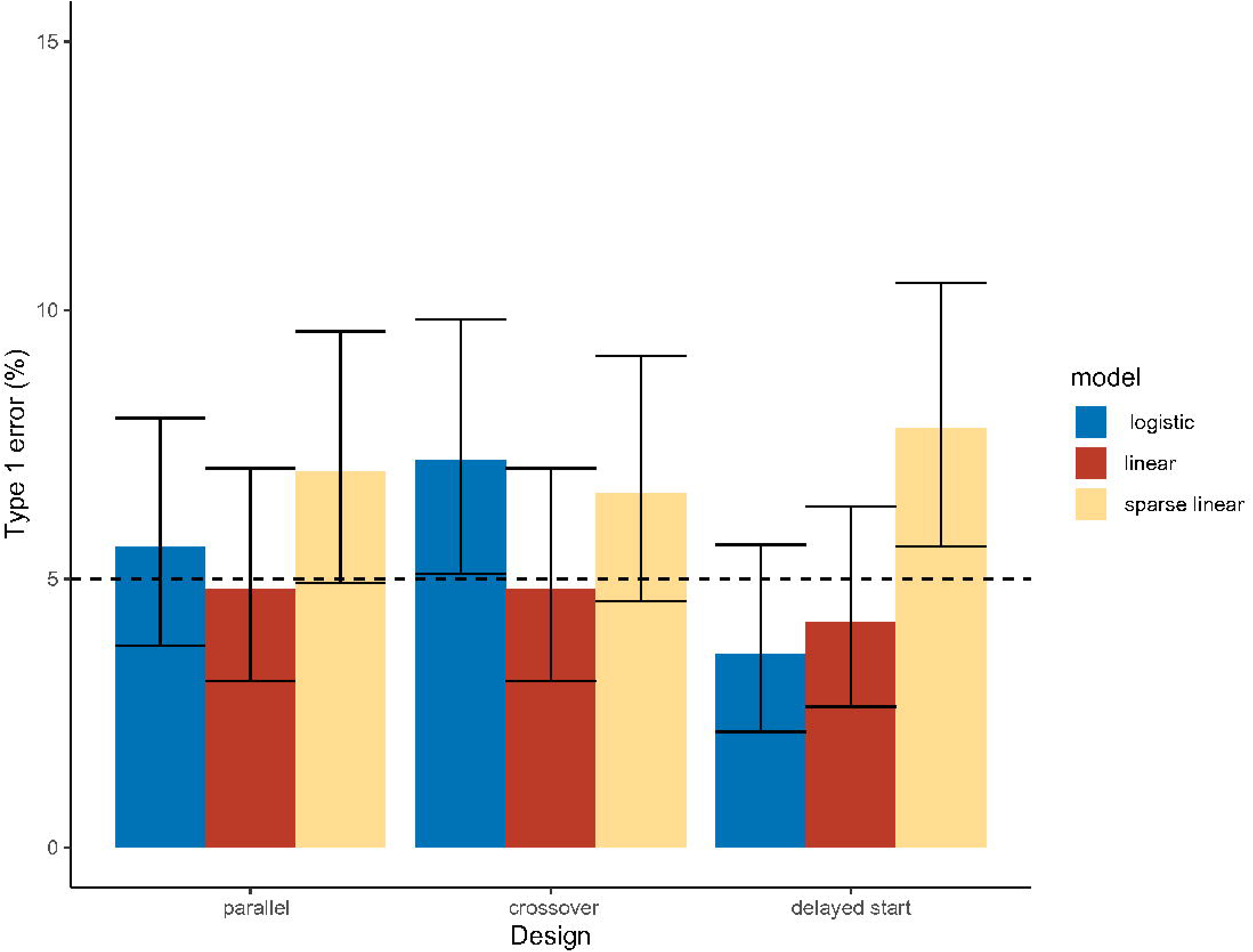
Type 1 error for the three designs and three analysis models on the slow progression scenario without a drug effect (DE=0%), N = 100 and assuming a trial duration of 5 years. The colours of the bars correspond to the model used for estimation

**Figure 5.**
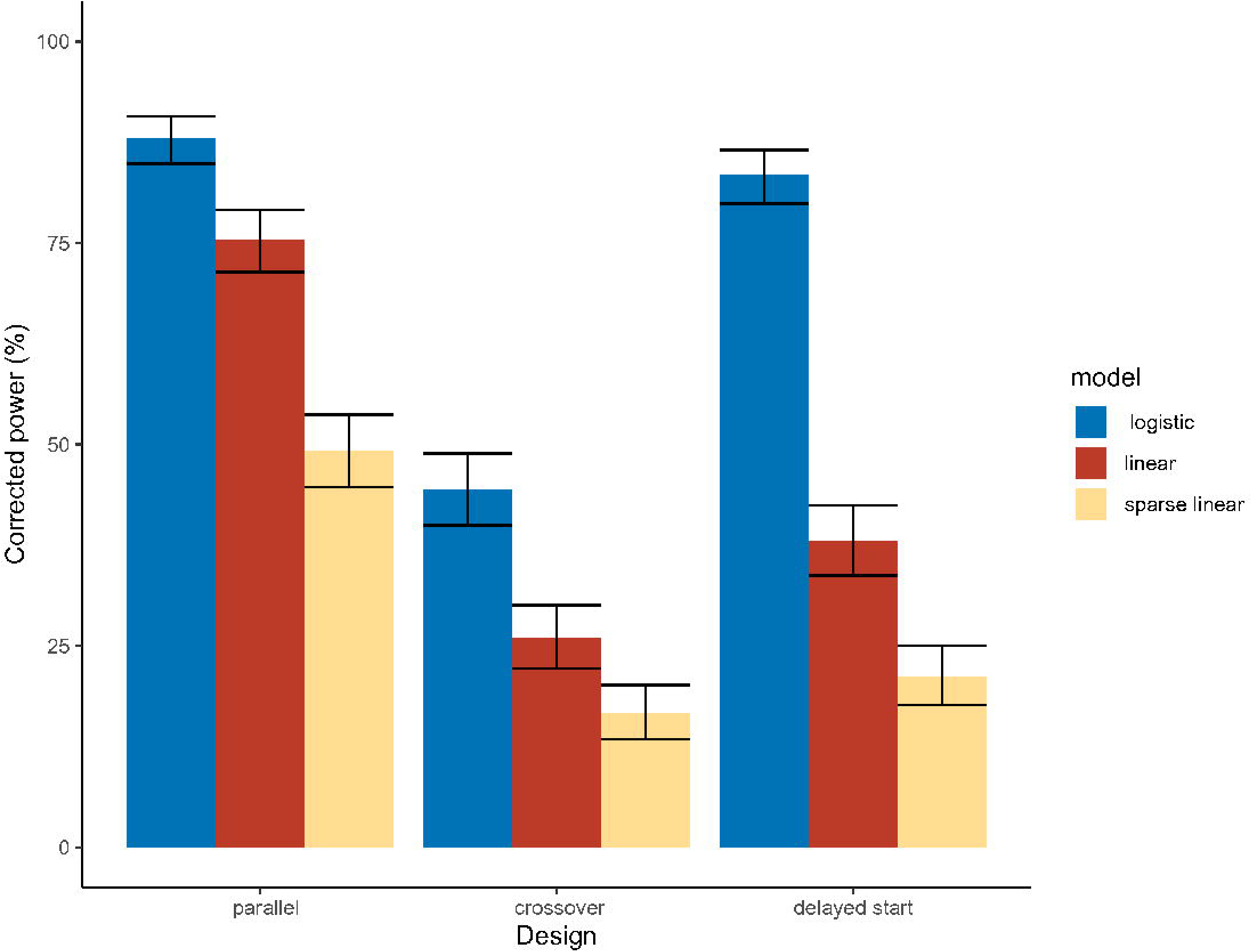
Corrected power for the three designs and three analysis models on the slow progression scenario assuming a drug effect DE=50%, N = 100 and a trial duration of 5 years. The colours of the bars correspond to the model used for estimation

**Figure 6.**
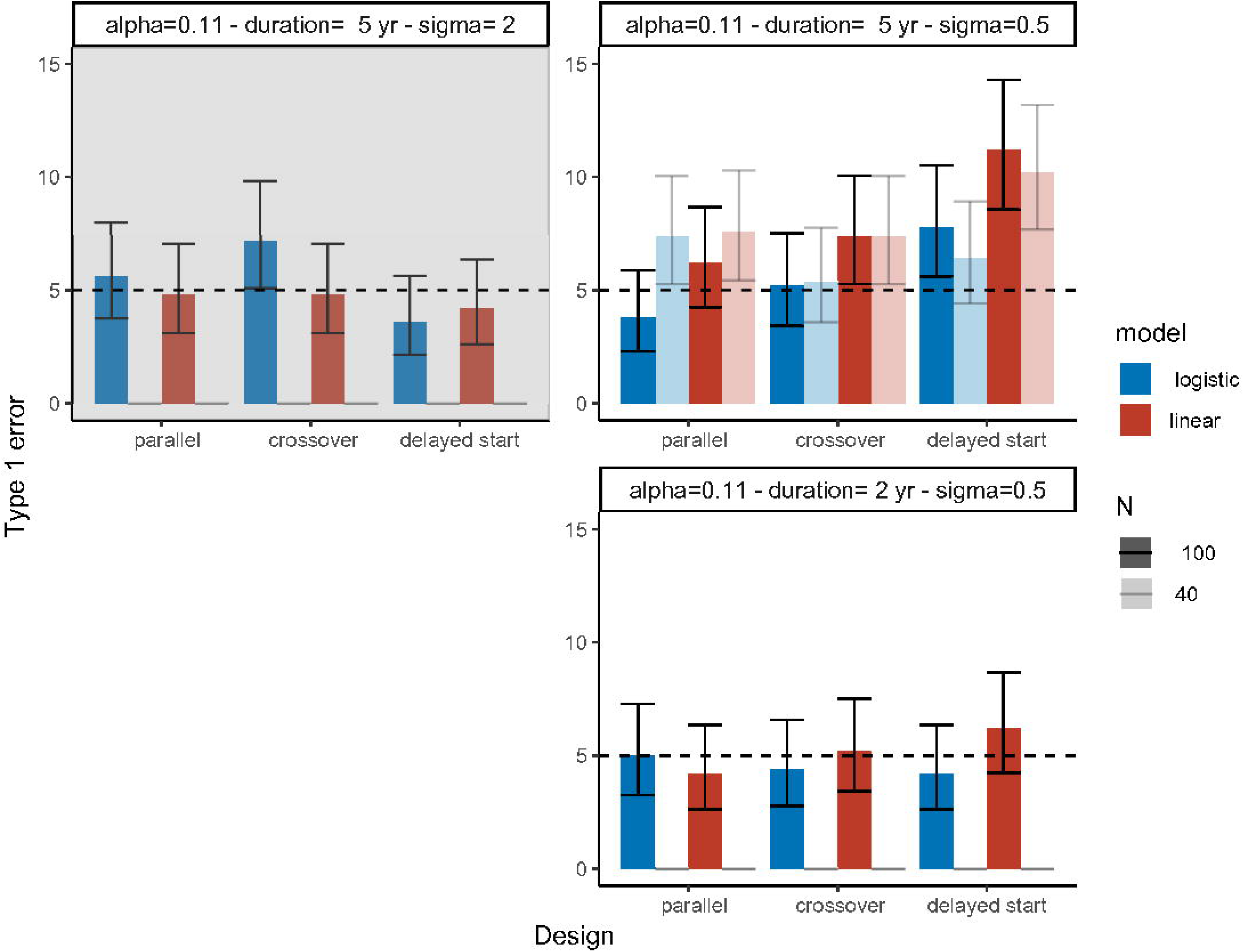
Type 1 error for the slow progression reference and the scenarios with duration = 5 years/ σ=0.5 and duration = 2 years/σ=0.5. The colours of the bars correspond to the model used for estimation. The upper left panel with a grey background highlights the reference scenario (slow progression). In the upper right panel, darker and lighter shades represent N=100 and N=40, respectively.

We then investigated the influence of a smaller magnitude of residual error (σ=0.5 and a duration 5 years) and the influence of a shorter trial duration (2 years and σ=0.5) on the type 1 error and corrected power. Type 1 error (Fig 5) was controlled in most scenarios for the logistic model and it was inflated in some scenarios for the linear model. In Figure 7, we show the corrected power for those scenarios. With a duration = 5 years and σ=0.5, the power was close to 100% for the logistic model in all designs. All designs were therefore re-evaluated with 40 patients to better compare the relative performance of each design. The corrected power is also close to 100% in almost all designs for the linear model. With a shorter duration of 2 years and a lower residual error, we observed powers similar to the slow progression reference scenario, with power going from 90% for the parallel and delayed designs to 57.2% for the cross-over design. In this scenario, the linear model had lower power than the logistic model, and the delayed start design performed as poorly as the cross-over design with power dropping by half compared to the parallel design.

**Figure 7.**
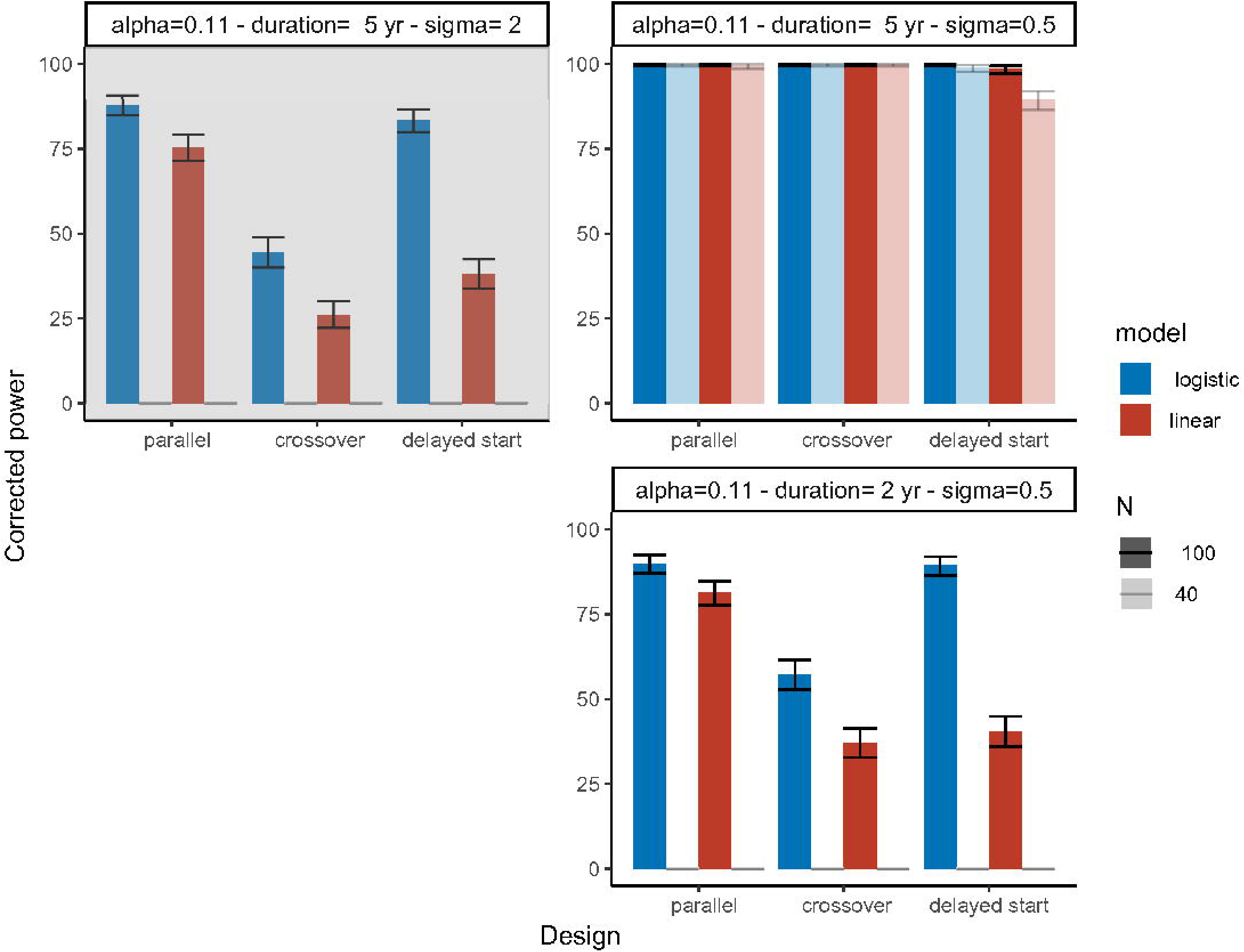
Corrected power for the slow progression reference and the scenarios with duration = 5 years/ σ=0.5 and duration = 2 years/σ=0.5. The colours of the bars correspond to the model used for estimation. The upper left panel with a grey background highlights the reference scenario (slow progression). In the upper right panel, darker and lighter shades represent N=100 and N=40 respectively

### 3.2 Simulation study: fast-progression case

In the fast progression reference scenario, the type 1 error was controlled for all designs for both the logistic and the linear models (Figure 8 bottom-left panel). The corrected powers (Figure 9 bottom-left panel), were similar to the slow progression reference case, with the highest power found for the parallel and delayed start designs, and a marked decrease for the crossover design. The corrected power for the linear model was much lower in all designs.

**Figure 8.**
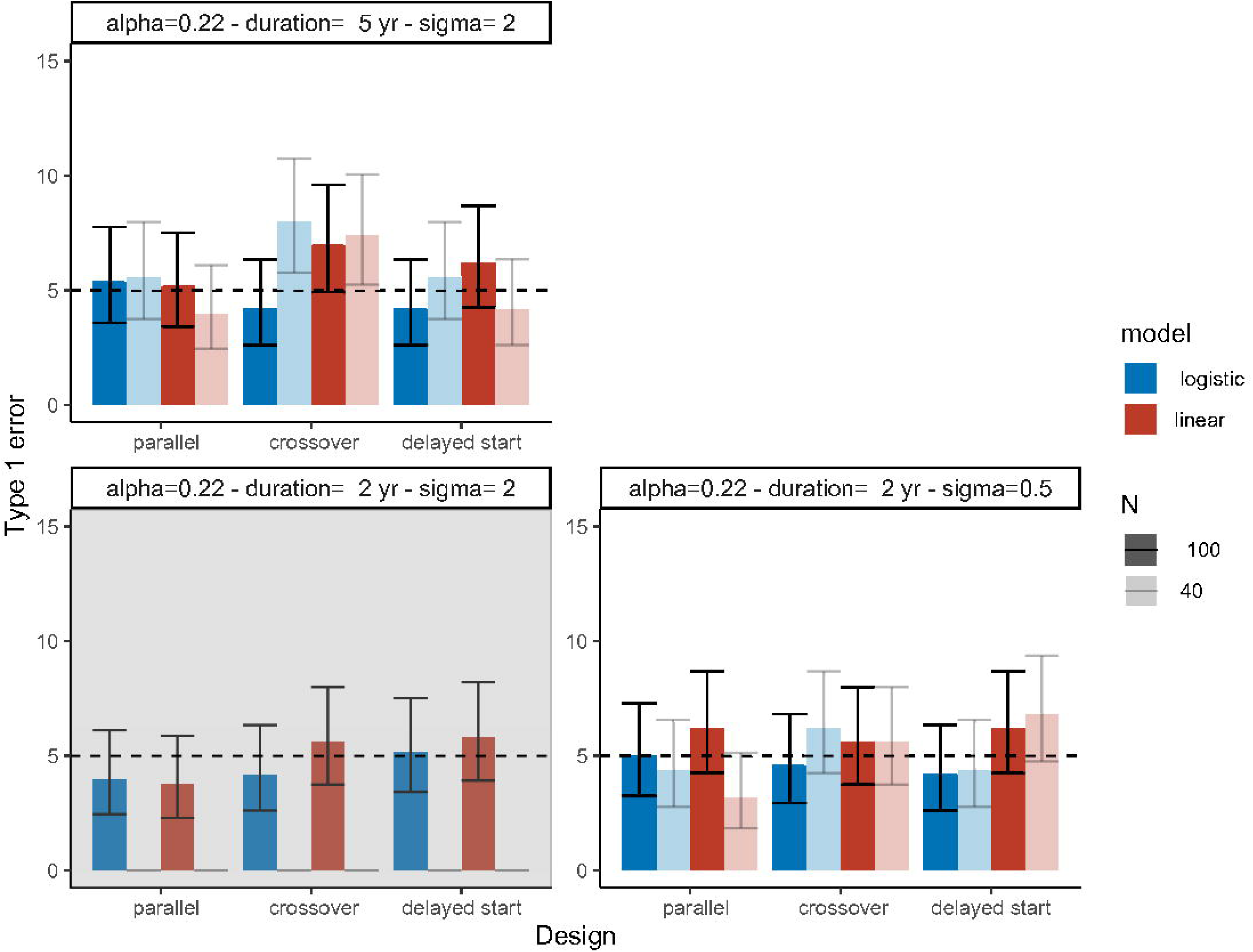
Type 1 error for the fast progression reference and the scenarios with duration = 5 years/ σ=2 and duration = 2 years/σ=0.5. The colours of the bars correspond to the model used for estimation. The upper left panel with a grey background highlights the reference scenario (fast progression). In the upper left and bottom right panels, darker and lighter shades represent N=100 and N=40 respectively.

**Figure 9.**
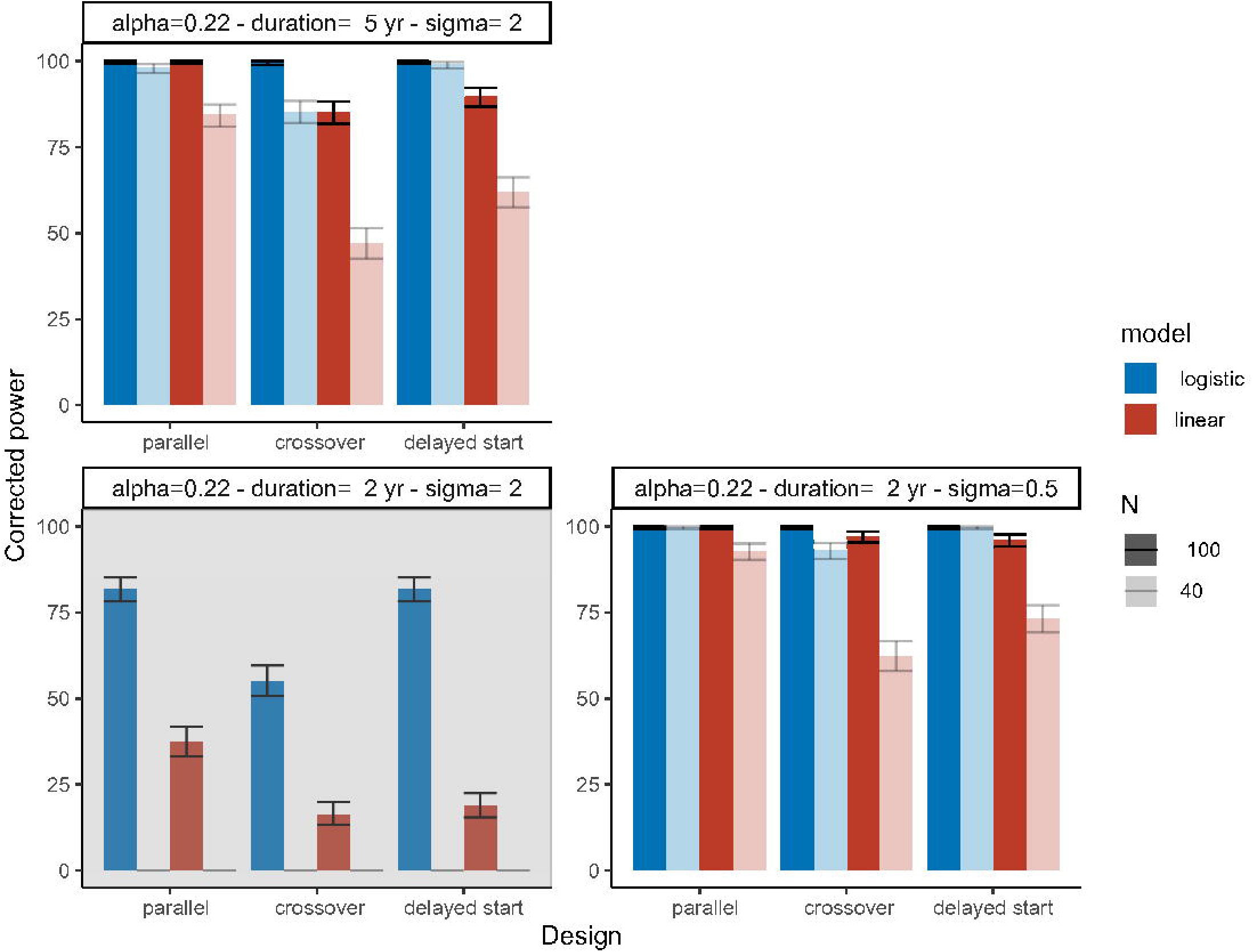
Corrected power for the fast progression reference and the scenarios with duration = 5 years/ σ=2 and duration = 2 years/σ=0.5. The colours of the bars correspond to the model used for estimation. The upper left panel with a grey background highlights the reference scenario (fast progression). In the upper left and bottom right panels, darker and lighter shades represent N=100 and N=40 respectively.

Varying the magnitude of residual error (σ=0.5) or considering a longer trial duration of trial did not impact type I error (Figure 8) which remained controlled in almost all scenarios for both the logistic and linear models. In Figure 9, we show the corrected power for those scenarios. For the logistic model, the power was close to 100% in all scenarios with 100 patients, and remained high with 40 patients for the parallel and delayed start design, decreasing by 5 to 15% for the crossover design. For the linear model, the corrected power was close to 100% with 100 patients when σ=0.5 for all 3 designs but was much lower with 40 patients than the logistic model.

## 4. Discussion

In this work, we performed a simulation study to evaluate the type 1 error and power to detect a treatment effect through longitudinal modelling for different designs, progression rates, durations and magnitudes of residual error. The settings for the simulation study were derived from a model describing the evolution of SARA scores in patients with ARCA^15^. We compared the modelling approach with an end-of-treatment analysis evaluating change from baseline.

In our settings, we used a sparse linear model to mimic an end-of treatment analysis, adding the mid trial point required for the crossover and delayed start designs to keep the same number of observations for all designs. The sparse linear model was outperformed by the logistic model. For the parallel design, using longitudinal modelling with non-linear mixed effect models enabled the power of the trial to go from 50% to 88% (and from 38% to 88% with a two-sample t-test with a change from baseline). The linear mixed effect model on the full data also performed worse than the logistic model, even when informing the base and slope parameters with the time since onset at inclusion in the study, although, in most scenarios, the power of the linear model was still relatively high when using a parallel design. This shows that part of the loss of power is attributable to model misspecification since data was simulated using a logistic model.

In this study, we found that, in both the slow and fast progression case, the parallel design performs the best, followed by the delayed start design which had almost identical power in all scenarios. The crossover design had lower power than the other two. The poor performance of the crossover design was unexpected, as, in some settings, such as bioequivalence studies, it was shown to have better performance in terms of power than a parallel design ^22^, due to the fact that it allows to identify treatment effect using each patient as its own control. This loss of power was not due to a bias in the estimation of the treatment effect as shown in Supplementary Figures S1-1 and S1-2. In our case, the underperformance of the crossover design could be explained by the fact that neurodegenerative ataxias are slowly progressive diseases. Even though using non-linear models accounts for the temporal and carry-over effect, the identification of the treatment effect relies on the observation of a progression under treatment. In the particular context of ARCAs we found (Hendrickx et al.) ^7^ that for the ARSACS population which we used as a basis for the slow-progression scenario, the change in SARA score was very progressive and variable depending on disease stage (from 0.05 to 1 point/year). The SARA score also increases especially slowly in the earlier stages, and here with uniform inclusion in the first 30 years after onset of symptom we expect SARA to progress by less than 0.25 points per year in half of the subjects included. This should also be contrasted with the high residual error (σ=2), implying that to identify a disease modifying treatment effect, it is important to observe the disease progression for a long period of time. In our simulation settings, the total duration was the same for all designs, so that disease progression was only followed for 2.5 years under treatment for each patient in the cross-over design. Furthermore, here the variability in progression rate is termed quite low (<30%), whereas the strength of a crossover trial is to differentiate between the variability in disease progression (a) and the treatment effect.

In our settings, the delayed start design performed almost as well as the parallel design. An advantage is that all patients eventually get treated, just like in single arm trial, while preserving the ability to identify untreated progression within the trial. A delayed start design was evaluated by Wang et al. ^12^ for its ability to detect a drug-induced disease modification in neurodegenerative disorders, as with a lasting effect on disease progression, patients who receive the treatment only in the second period won’t catch up to the patients who received the treatment earlier, resulting in parallel curves between the two treatment groups. We believe that, in our settings, the delayed start design outperforms the crossover design because in one group, patients are observed under treatment for the full duration of the trial, unlike the crossover design.

In this work, we based our simulation settings on natural disease progression in the ARSACS population of the ARCA registry assuming a disease modifying treatment effect on disease progression. Even though there are currently no disease-modifying treatments for most ARCAs, several clinical trials are ongoing for molecular therapies or gene therapies. For example, FDA just approved the use of levacetylleucine for Niemann-Pick disease type C ^10^ (NPC). It here followed a clinical trial protocol ^23^ ^24^ that will evaluate the efficacy of the drug also – in addition to NPC -on two other ultra-rare neurodegenerative diseases, including Ataxia Telangiectasia, an ARCA with fast progression ^25^. They evaluated the efficacy of the compound for Niemann-Pick disease type C with 60 patients, with a crossover design including two 12-weeks periods using the SARA score as the primary endpoint. The authors were able to show evidence of efficacy, however, they also emphasized that the proposed design could only demonstrate the symptomatic effect of the treatment and that they would need a larger sample size and a longer trial to show an effect on disease progression. In their protocol, they included a single-arm open label extension where they would monitor the progression for two years under treatment to investigate the effect on the disease progression. This difference of assumption on treatment effect (symptomatic versus disease-modifying) can explain why they could show a treatment effect with a crossover trial while, in our simulations, the crossover trial could not reach such levels of power, even with a duration of two years. The trial aiming to evaluate the efficacy of the compound on Ataxia Telangiectasia is a single arm trial ^26^, where patients are monitored 6 weeks with treatment and also for a 6-week post-treatment washout period. There are currently 39 patients enrolled (patients of 6 years of age and older). The primary outcome measured is the Clinical Impression of Change in Severity (CI-CS) (7-point Likert scale, from −3=significantly worse to +3= significantly improved) and the SARA score is measured as a secondary outcome.

Ataxia Telangiectasia could be compared to our fast progression reference. Again, the trial duration is much shorter but the assumption on the magnitude of treatment effect is different. Bruegermann et al. ^27^ monitor the evolution of the SARA score on 6 patients with Ataxia Telangiectasia with the same compound. They found that, after one month of treatment, the SARA score is significantly lower (by 4 points on average), and that the SARA score goes back up after patients stop the treatment, suggesting the effect could be symptomatic.

However, this trial design confirms that it would be feasible (if these effect sizes can indeed be independently confirmed) to enroll 40 patients in the fast-progression case. In addition to molecular therapies, clinical trials investigating the efficacy of gene therapy are also being conducted in ARCAs. In fact, the three ongoing gene therapy clinical trials in Friedrich’s Ataxia ^28^ ^29^ ^30^, will follow patients for 5 years, with a disease progression in this genetic diagnostic similar to that of ARSACS ^14^, but with much lower sample sizes, as those are phase 1 trials.

The parameters we used in the slow progression scenario correspond to the estimates found when modelling the change in SARA score in the ARSACS patients of the ARCA registry ^7^, where we found a high variability of the SARA score ( σ=2) in comparison to the yearly rate of disease progression. The implication of the poor signal/noise ratio as we saw in our simulations is the longer trial duration needed to discriminate natural progression from slower progression under treatment. Reducing the variability in the SARA score would open the possibility for shorter trials, as shown by the simulations performed with σ=0.5. Grobe-Einsler et al. ^31^, evaluated the property of repeated measures of the score. They selected 5 subitems of the score which can be videotaped by the patient themselves without requiring an on-site examiner (excluding the sitting and two fine motor movement subitems), and evaluated the score on 12 patients with ataxia twice a day for 14 days. They notably highlighted that there was no statistically significant training effect over the two-week period and that there was a four-fold reduction of variance of residual error by averaging all SARA assessments. This could confirm that the high residual variability found in our model is inherent to the SARA score and that repeating measures could improve the assessment of an individual’s SARA score, even though this could be difficult to conduct in practice. In our simulation framework, we tested scenarios with a lower magnitude of residual error. We could assume that such a scenario would be feasible by either reporting an average SARA score over several measurements or increasing the visit frequency. Additionally, in our framework, for simulation and estimation the SARA score (which ranges from 0 to 40 points) was treated as continuous. We could also expect a small drop in power in all scenarios if the simulated score was discretized before the estimation. Alternatively, another modelling approach could be applied, with an Item Response Theory (IRT) model for example ^14,32^, which would keep the discrete nature of the data and which would leverage information from each sub item of the SARA score instead of the total score.

In the simulation framework, we chose the SARA score as the primary endpoint to assess treatment efficacy. SARA score is the currently most widely used outcome to assess disease severity in ataxia patients. We chose the SARA score because it is included in most clinical trials, at least as a secondary endpoint or used in the inclusion criterion. It is a composite score comprised of 8 subitems evaluating gait, stance, sitting, speech, and 4 other items for fine motor movements. It was shown in Weyer al.^33^ to be a reliable and valid measure is disease severity in patients with ataxia, but it was weakly correlated to disease duration (time since onset of symptoms). Furthermore, the FDA is also investigating a modified version of the SARA score, called the modified functional Scale for Assessment and Rating of Ataxia (f-SARA). In fact, Potashman et al. ^34^ discuss that the score was modified to only include the gait, stance, speech and sitting items, because, in the context of a clinical trial that would last one year, the other items were considered not sensitive enough to capture change in disease progression. Therefore, even though, in patient registries, the SARA score would be more meaningful to capture long term disease progression- and also upper limb functioning, the f-SARA score could be more relevant in the context of a clinical trial with a shorter duration, where the score might be more sensitive to change-although it would lose informational value, as shown by our recent IRT models of the SARA ^14,32^. Finally, for ARCAs, composite clinical scores are being investigated because there are currently no validated biomarkers that can be applied to all genotypes. However, other outcomes are being investigated, such as quantitative motor assessment, called digital biomarkers ^35^. The goal is to quantify variability and stability of patient movement using sensors. They are being developed and being validated as a quantitative and continuous metrics that could be used as an outcome in clinical trials ^36^. Fluid biomarkers are also being investigated, such as the Neurofilament Light chain (Nfl), a biomarker of neurodegeneration. In fact, Donath et al. ^37^ showed that Nfl levels were significantly higher in Ataxia Telangiectasia patients than healthy individuals. It was also shown in Li et al. ^38^ that, for patients with Spinocerebellar ataxia type 3, serum levels of Nfl were significantly correlated with disease severity (with both the ICARS and SARA scores). This biomarker is also included as a secondary outcome in a clinical trial ^39^ that evaluates the effect of a compound on patients with Ataxia Telangiectasia. Given the fact that, in our simulation framework, we assume that the SARA score is continuous, our modelling approach would be compatible with such continuous biomarkers.

In our work, the simulation framework for clinical trial designs was built upon a previously built non-linear mixed effect model ^7^, and several assumptions were made. The four-parameter logistic model was built using data from the ARCA registry ^15^, a multicentric registry of patients with Autosomal Recessive Cerebellar Ataxia. Then, since there is currently no disease-modifying drug approved for most ARCAs, we hypothesised that the treatment would reduce the a parameter, which implies that the treatment would have an effect on both the disease progression rate and the inflection point of the logistic curve. We could have implemented treatment effect differently, for example as a change in f3, which controls the inflexion point, making patients progressing later, or in y, assuming that the treatment would decrease the maximum score reachable by a patient, or a combination of both. Also, as discussed by Bermova-Ertl et al. ^10^, it is possible that a disease modifying treatment could also have a symptomatic effect, meaning that, for the patients, the SARA score could also decrease temporarily. Then, we also made an assumption on the inclusion criterion of the trials. The range of 0 to 30 years since onset of the disease was chosen because, in the slow progression case, it would translate to a range of 0-20 points of SARA score and, according to Benatar et al ^40^, patients with a neurodegenerative disease would benefit from a clinical trial for a disease modifying drug when they are at most at a mild stage of the disease. Even though recruiting patients during the later stages of the range (20-30 years) would yield higher power ^7,14^, an inclusion criterion based on an heterogeneous time since onset at inclusion was chosen because it would be the most realistic possibility given the rare nature of the disease, which was confirmed by many of the ongoing clinical trials presented above, some of which included patients starting at 6 years of age. However, since this range was chosen for the slow progression case, it might not be completely comparable for the fast progression case (on average, patients would reach a SARA score of 20 points 15 years after the onset of symptoms). An important assumption in model-based analyses is that the model is well specified, meaning, in our case, that the logistic model is the simulation model describing the evolution of the SARA score. In our simulation framework, the logistic model was therefore advantaged compared the linear and sparse linear models, even though the parameters of the logistic model were re-estimated. While a four-parameter logistic model is a flexible and generic model, other, more robust, approaches could have been used to handle model misspecification, such as model averaging ^3,41,42^. Finally, to identify a treatment effect, we used the likelihood ratio test, which relies on an asymptotic assumption. In non-linear mixed effect models, it translates to having sufficient patients and observations per patient. In our simulations, we considered that this condition was met with 40 and 100 patients, with 6-11 observations per patients. However, if we wanted to assess smaller trial populations (<20), we would need to use other statistical tests that do not rely on that asymptotic assumption.

## 5. Conclusion

In this work, we proposed a simulation framework to compare clinical trial designs and settings in the context of rare neurological diseases, using as showcase basis for the simulations the disease group of ARCAs, characterised by small sample sizes (less than 100), patient heterogeneity (in terms of disease severity at inclusion), use of real-world data, lack of curative/efficient treatments. We studied the influence of using longitudinal models versus more standard statistical analyses, the influence of disease progression rate, trial duration and magnitude of residual error on the type 1 error and power on a clinical trial that would quantify a disease-modifying treatment effect. In our settings the parallel performed best. The delayed start design had similar performance with the advantage that all patients are treated in this design. In our framework, the crossover design had lower power, mainly due to the slow disease progression coupled with high residual error, requiring to observe disease progression for a longer time (at least in a disease-modifying treatment setting, as tested here). Reducing the sample size to match disease rarity would require smaller residual variability in the primary outcome, here the SARA score, to better identify disease progression, which would need to be achieved by less noisy endpoints or more frequent assessments.

## Data Availability

The code for the simulation used in the study is available in a repository

https://doi.org/10.5281/zenodo.14755801

## Acknowledgments

This work was funded by the European Joint Programme on Rare Diseases (EJP RD) Joint Transnational Call 2019 for the EJP RD WP20 Innovation Statistics consortium “EVIDENCE-RND” focusing on “Innovative Statistical Methodologies to Improve Rare Diseases Clinical Trials in Limited Populations” under the EJP RD Grant Agreement (n°825575) (to M.K, R.S. and M.S.); as well as by the European Union, project European Rare Disease Research Alliance (ERDERA), GA n°101156595, funded under call HORIZON-HLTH-2023-DISEASE-07 (to M.S. R.S, and F.M.). Moreover, work in this project was supported by the Clinician Scientist programme “PRECISE.net” funded by the Else Kröner-Fresenius-Stiftung (to M.S., R.S. and A.T) and the Bundesministerium für Bildung und Forschung (BMBF) through funding for the TreatHSP network (grant 01GM2209A to R.S.). This work was supported by members of the Evidence-RND consortium, which includes Xiaomei Chen, Nicole Maria Heussen, Ralf-Dieter Hilgers, Thomas Klockgether, Yevgen Ryeznik, Oleksandr Sverdlov. R.S. and M.S. are members of the European Reference Network for Rare Neurological Diseases – Project ID 739510.

## Ethical approval and informed consent statements

For this study, we used a previously published model to simulate data, so no personal data was collected and/or processed.

## Bibliography

1. Research C for DE and. E9 Statistical Principles for Clinical Trials, https://www.fda.gov/regulatory-information/search-fda-guidance-documents/e9-statistical-principles-clinical-trials (2020, accessed 18 December 2024).

2. Research C for DE and. Real-World Data: Assessing Registries To Support Regulatory Decision-Making for Drug and Biological Products, https://www.fda.gov/regulatory-information/search-fda-guidance-documents/real-world-data-assessing-registries-support-regulatory-decision-making-drug-and-biological-products (2023, accessed 17 December 2024).

3. Buatois S, Ueckert S, Frey N, et al. cLRT-Mod: An efficient methodology for pharmacometric model-based analysis of longitudinal phase II dose finding studies under model uncertainty. Statistics in Medicine 2021; 40: 2435–2451.

4. Karlsson KE, Vong C, Bergstrand M, et al. Comparisons of Analysis Methods for Proof-of-Concept Trials. CPT Pharmacometrics Syst Pharmacol 2013; 2: e23.

5. Synofzik M, Puccio H, Mochel F, et al. Autosomal Recessive Cerebellar Ataxias: Paving the Way toward Targeted Molecular Therapies. Neuron 2019; 101: 560–583.

6. Synofzik M, van Roon-Mom WMC, Marckmann G, et al. Preparing n-of-1 Antisense Oligonucleotide Treatments for Rare Neurological Diseases in Europe: Genetic, Regulatory, and Ethical Perspectives. Nucleic Acid Ther 2022; 32: 83–94.

7. Hendrickx N, Mentré F, Traschütz A, et al. Prediction of Individual Disease Progression Including Parameter Uncertainty in Rare Neurodegenerative Diseases: The Example of Autosomal-Recessive Spastic Ataxia Charlevoix Saguenay (ARSACS). AAPS J 2024; 26: 57.

8. Traschütz A, Adarmes-Gómez AD, Anheim M, et al. Responsiveness of the Scale for the Assessment and Rating of Ataxia and Natural History in 884 Recessive and Early Onset Ataxia Patients. Annals of Neurology; n/a. DOI: 10.1002/ana.26712.

9. Traschütz A, Adarmes-Gomez AD, Anheim M, et al. Autosomal Recessive Cerebellar Ataxias in Europe: Frequency, Onset, and Severity in 677 Patients. Movement Disorders 2023; 38: 1109–1112.

10. Bremova-Ertl T, Ramaswami U, Brands M, et al. Trial of N-Acetyl-l-Leucine in Niemann–Pick Disease Type C. New England Journal of Medicine 2024; 390: 421–431.

11. Liu-Seifert H, Andersen SW, Lipkovich I, et al. A Novel Approach to Delayed-Start Analyses for Demonstrating Disease-Modifying Effects in Alzheimer’s Disease. PLoS ONE 2015; 10: e0119632.

12. Wang D, Robieson W, Zhao J, et al. Statistical considerations in a delayed-start design to demonstrate disease modification effect in neurodegenerative disorders. Pharmaceutical Statistics 2019; 18: 407–419.

13. Schmitz-Hübsch T, du Montcel ST, Baliko L, et al. Scale for the assessment and rating of ataxia: Development of a new clinical scale. Neurology 2006; 66: 1717–1720.

14. Hamdan A, Hendrickx N, Hooker AC, et al. Longitudinal Analysis of Natural History Progression of Rare and Ultra-Rare Cerebellar Ataxias Using Item Response Theory. Clinical Pharmacology & Therapeutics; n/a. DOI: 10.1002/cpt.3466.

15. Traschütz A, Reich S, Adarmes AD, et al. The ARCA Registry: A Collaborative Global Platform for Advancing Trial Readiness in Autosomal Recessive Cerebellar Ataxias. Front Neurol 2021; 12: 677551.

16. Synofzik M, Soehn AS, Gburek-Augustat J, et al. Autosomal recessive spastic ataxia of Charlevoix Saguenay (ARSACS): expanding the genetic, clinical and imaging spectrum. Orphanet Journal of Rare Diseases 2013; 8: 41.

17. Beal S, Sheiner LB, Boeckmann A, et al. NONMEM User’s guides. (1989-2009).

18. Lindbom L, Pihlgren P, Jonsson EN. PsN-Toolkit--a collection of computer intensive statistical methods for non-linear mixed effect modeling using NONMEM. Comput Methods Programs Biomed 2005; 79: 241–257.

19. Delyon B, Lavielle M, Moulines E. Convergence of a stochastic approximation version of the EM algorithm. The Annals of Statistics 1999; 27: 94–128.

20. Wang Y. Derivation of various NONMEM estimation methods. J Pharmacokinet Pharmacodyn 2007; 34: 575–593.

21. 21. R Core Team. R: A language and environment for statistical computing, https://www.R-project.org/ (2022).

22. Lathyris DN, Trikalinos TA, Ioannidis JP. Evidence from crossover trials: empirical evaluation and comparison against parallel arm trials. International Journal of Epidemiology 2007; 36: 422–430.

23. Fields T, Patterson M, Bremova-Ertl T, et al. A master protocol to investigate a novel therapy acetyl-l-leucine for three ultra-rare neurodegenerative diseases: Niemann-Pick type C, the GM2 gangliosidoses, and ataxia telangiectasia. Trials 2021; 22: 84.

24. IntraBio Inc. Effects of N-Acetyl-L-Leucine on Niemann-Pick Disease Type C (NPC): A Phase III, Randomized, Placebo-controlled, Double-blind, Crossover Study. Clinical Trial Registration NCT05163288, clinicaltrials.gov, https://clinicaltrials.gov/study/NCT05163288 (11 September 2023, accessed 26 November 2024).

25. Petley E, Yule A, Alexander S, et al. The natural history of ataxia-telangiectasia (A-T): A systematic review. PLoS ONE 2022; 17: e0264177.

26. IntraBio Inc. Effects of N-Acetyl-L-Leucine on Ataxia-Telangiectasia (A-T): A Multinational, Multicenter, Open-label, Rater-blinded Phase II Study. Clinical Trial Registration NCT03759678, clinicaltrials.gov, https://clinicaltrials.gov/study/NCT03759678 (29 August 2023, accessed 26 November 2024).

27. Brueggemann A, Bicvic A, Goeldlin M, et al. Effects of Acetyl-DL-Leucine on Ataxia and Downbeat-Nystagmus in Six Patients With Ataxia Telangiectasia. J Child Neurol 2022; 37: 20–27.

28. Weill Medical College of Cornell University. Phase IA and IB Study of AAVrh.10hFXN Gene Therapy for the Cardiomyopathy of Friedreich&#x27;s Ataxia. Clinical Trial Registration NCT05302271, clinicaltrials.gov, https://clinicaltrials.gov/study/NCT05302271 (15 October 2024, accessed 26 November 2024).

29. Astellas Gene Therapies. A Multicenter, Open-Label, Dose Escalation, Phase 1b Study to Evaluate the Safety, Tolerability and Preliminary Efficacy of ASP2016 for Friedreich Ataxia Cardiomyopathy. Clinical Trial Registration NCT06483802, clinicaltrials.gov, https://clinicaltrials.gov/study/NCT06483802 (19 November 2024, accessed 26 November 2024).

30. Lexeo Therapeutics. A Phase 1&#x2F;2 Study of the Safety and Efficacy of LX2006 Gene Therapy in Participants With Cardiomyopathy Associated With Friedreich&#x27;s Ataxia. Clinical Trial Registration NCT05445323, clinicaltrials.gov, https://clinicaltrials.gov/study/NCT05445323 (13 November 2024, accessed 26 November 2024).

31. Grobe-Einsler M, Taheri Amin A, Faber J, et al. Development of SARAhome, a New Video-Based Tool for the Assessment of Ataxia at Home. Mov Disord 2021; 36: 1242– 1246.

32. Hamdan A, Hooker AC, Chen X, et al. Item performance of the scale for the assessment and rating of ataxia in rare and ultra-rare genetic ataxias. CPT: Pharmacometrics & Systems Pharmacology 2024; 13: 1327–1340.

33. Weyer A, Abele M, Schmitz-Hübsch T, et al. Reliability and validity of the scale for the assessment and rating of ataxia: a study in 64 ataxia patients. Mov Disord 2007; 22: 1633–1637.

34. Potashman M, Rudell K, Pavisic I, et al. Content Validity of the Modified Functional Scale for the Assessment and Rating of Ataxia (f-SARA) Instrument in Spinocerebellar Ataxia. Cerebellum 2024; 23: 2012–2027.

35. Hermle D, Schubert R, Barallon P, et al. Multifeature quantitative motor assessment of upper limb ataxia including drawing and reaching. Annals of Clinical and Translational Neurology 2024; 11: 1097.

36. Ilg W, Müller B, Faber J, et al. Digital Gait Biomarkers Allow to Capture 1-Year Longitudinal Change in Spinocerebellar Ataxia Type 3. Mov Disord 2022; 37: 2295–2301.

37. Donath H, Woelke S, Schubert R, et al. Neurofilament Light Chain Is a Biomarker of Neurodegeneration in Ataxia Telangiectasia. *Cerebellum (London*, England) 2021; 21: 39.

38. Li Q-F, Dong Y, Yang L, et al. Neurofilament light chain is a promising serum biomarker in spinocerebellar ataxia type 3. Mol Neurodegener 2019; 14: 39.

39. The University of Queensland. Single Arm Open-label Clinical Trial in Ataxia-telangiectasia to Test the Effects of Nicotinamide Riboside on Ataxia Scales, Immune Function, and Neurofilament Light Chain. Clinical Trial Registration NCT06324877, clinicaltrials.gov, https://clinicaltrials.gov/study/NCT06324877 (16 April 2024, accessed 26 November 2024).

40. Benatar M, Wuu J, McHutchison C, et al. Preventing amyotrophic lateral sclerosis: insights from pre-symptomatic neurodegenerative diseases. Brain 2022; 145: 27–44.

41. Philipp M, Tessier A, Donnelly M, et al. Model-based bioequivalence approach for sparse pharmacokinetic bioequivalence studies: Model selection or model averaging? Statistics in Medicine 2024; 43: 3403–3416.

42. Buatois S, Ueckert S, Frey N, et al. Comparison of Model Averaging and Model Selection in Dose Finding Trials Analyzed by Nonlinear Mixed Effect Models. AAPS J 2018; 20: 56.

