## Supplementary Material for "Comparing randomized trial designs to estimate treatment effect in rare diseases with longitudinal models: a simulation study showcased by Autosomal Recessive Cerebellar Ataxias using the SARA score"

(1) Université Paris Cité, IAME, Inserm, F-75018, Paris, France
(2) Pharmacometrics Research Group, Department of Pharmacy, Uppsala University, Uppsala, Sweden
(3) Division Translational Genomics of Neurodegenerative Diseases, Hertie Institute for Clinical Brain Research (HIH), University of Tübingen, Tübingen, Germany
(4) German Center for Neurodegenerative Diseases (DZNE), Tübingen, Germany.
(5) Centre de Recherche du CHUS Et du Centre de Santé Et Des Services Sociaux du Saguenay-Lac-St-Jean, Faculté de Médecine, Université de Sherbrooke, Québec, Canada.
(6) Division of Neurodegenerative Diseases, Department of Neurology, Heidelberg University Hospital, Germany
(7) Univ Rennes, Inserm, EHESP, Irset - UMR_S 1085, 35000, Rennes, France.

**Supplementary material**

In this section we show the distribution of Estimation Errors (EE) of the Drug Effect parameter for the two reference scenarios (slow progression and fast progression) when estimated with the true (logistic) model including DE, under H0 (DE=0) for the type 1 error and H1 (DE=0.5) for the power. For replicate i, we have

$$EE_{i}=\hat{DE_{i}}-DE_{sim}$$

With $\hat{DE_{i}}$ the estimated drug effect in the i^th^ replicate, and $DE_{sim}$ the drug effect used for the simulation (0 or 0.5).

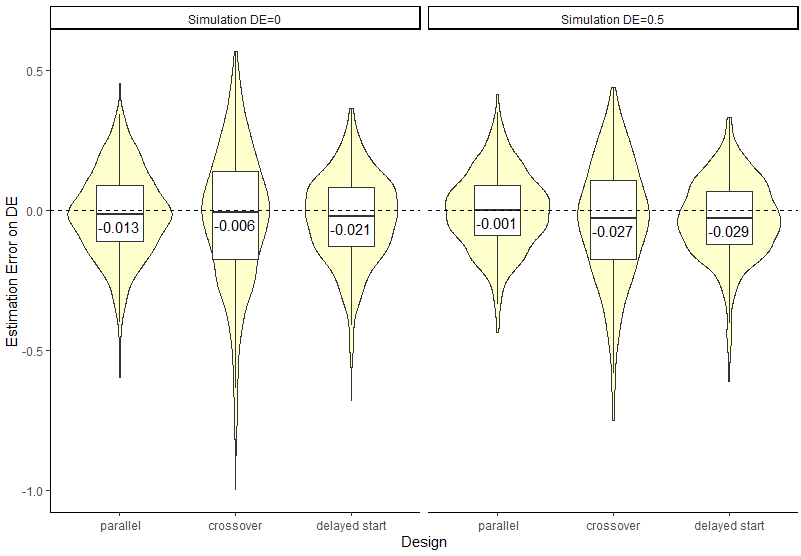

**Figure S1-1**. Violin plot of the Estimation Error on DE for each design for the **slow progression** case, with a simulated drug effect of 0 (left panel) or 0.5 (right panel)
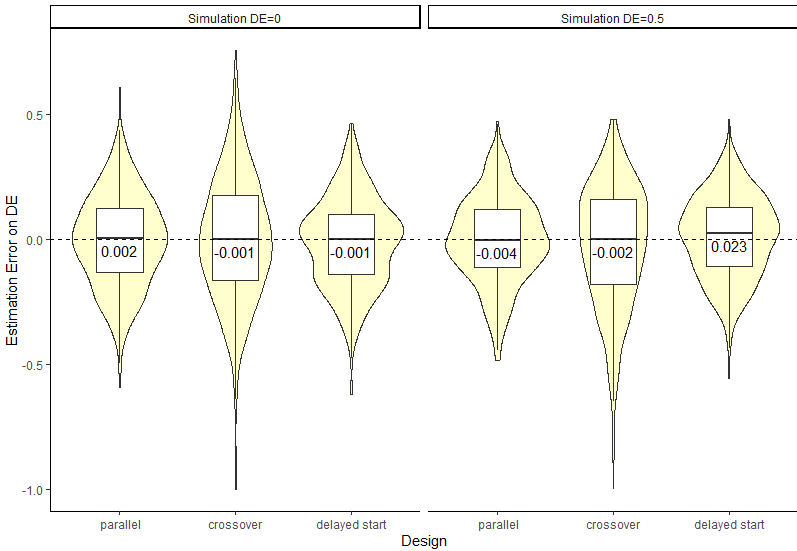

**Figure S1-2**. Violin plot of the Estimation Error on DE for each design for the **fast progression** case, with a simulated drug effect of 0 (left panel) or 0.5 (right panel)
